# Virtual reality based mindfulness applications: a commercial health app review

**DOI:** 10.1101/2025.03.21.25324405

**Authors:** Shraboni Ghosal, Mengying Zhang, Angeliki Bogosian, Elizabeth Marsh, Trudi Edginton, Emma Stanmore, Siobhan O’Connor

## Abstract

**Introduction:** Mindfulness can positively impact physical and mental health, but face-to-face programmes are limited by poor accessibility, availability, and cost. Virtual reality (VR) offers immersive audio-visual environments that could improve mindfulness practice.

**Aim:** To evaluate commercially available VR apps related to mindfulness.

**Methods:** App stores and relevant online platforms were searched for VR apps related to mindfulness. Results were screened against eligibility criteria and relevant data extracted. Six raters used the Mobile Application Rating Scale (MARS) to assess the quality of VR apps.

**Results:** Five VR apps related to mindfulness were included i.e., Headspace XR, Hoame, Innerworld, Maloka and TRIPP. These provided access to meditative and mindfulness sessions, guided by virtual instructors in some cases, and situated in a range of virtual landscapes accompanied by sound or music. TRIPP received the highest average MARS score (4.1), followed by Hoame (3.8), Maloka (3.6), Headspace XR (3.4) and Innerworld (3.3). Most VR apps scored the highest on functionality (3.4 to 4.2), while the information category scored the lowest (3.1 to 3.7). The intra-class correlation was moderate.

**Conclusion:** This review provides important insights into VR apps related to mindfulness such as their availability and quality. Only five VR apps were identified related to mindfulness practice with an overall moderate MARS quality score (3.62/5.00). These may provide a convenient and immersive way to access and engage in regular mindfulness practice, particularly for novices. Rigorous scientific research should assess the effectiveness of these VR apps in improving physical and mental health through immersive mindfulness practice.

## Introduction

Mindfulness is a course or state of consciousness, by which one can be aware of emotions, feelings, beliefs, and understandings in the present moment without making judgments ^1^. With its roots in Buddhism, it is a psychological attribute that can help a person focus on the present moment without being critical. Mindfulness incorporates breathing or relaxation techniques that may reduce stress, depression, anxiety, psychological distress, blood pressure and resting heart rate ^2^. Mindfulness based stress reduction and mindfulness-based cognitive therapy are standardised and commonly available Mindfulness Based Programmes (MBPs) which incorporate contemplative practices such as breathing, sitting, walking or meditation, understanding experiences, and letting go of long-term avoidance of experiences ^3,4^. Randomised controlled trials have shown significant improvements in mental health and wellbeing (i.e., anxiety, depression, psychological distress), acceptance, compassion, non-attachment ^5,6^, and quality of life and mood from MBPs ^7^. Several systematic reviews have shown mindfulness can be effective in clinical and non-clinical populations as it can improve psychological functioning by increasing self-awareness, improving emotional balance, and coping skills ^8,9^. However, face-to-face MBPs can suffer from limited accessibility and availability, and cost, with some people less comfortable with in person group-based mindfulness practice ^10,11^.

Digital health technologies offer a way to overcome some of the constraints of face-to-face MBPs. A review of mobile mindfulness applications (apps) found a mix of apps (n=23) providing videos and text to explain mindfulness with others relying upon guided meditation tracks to educate users ^12^. However, few had high ratings on the Mobile App Rating Scale (MARS) in terms of their visual design, level of engagement, functionality, and information quality ^12^. In addition, the review concluded that many apps claimed to be mindfulness-related but most only offered guided meditation, timers, or reminders. Regardless, a recent systematic review and meta-analysis of 45 trials of mindfulness apps reported only small improvements in anxiety and depression ^13^. However, a review of mindfulness apps in European app stores showed they are often of low quality, lack data security, have no privacy policy, or are not based on mindfulness or behaviour change techniques ^14^. Furthermore, any smartphone app is limited in terms of the level of interaction and visualisations it can provide.

Virtual reality (VR) is a rapidly advancing technology where users are immersed in three-dimensional audio-visual environments via a wraparound VR headset and hand controllers or haptic gloves. VR can be used to simulate real-life environments and situations as well as more abstract worlds where users can interact with objects while navigating around a virtual space filled with sounds or music. It is increasingly being used in healthcare for managing chronic conditions such as anxiety, depression, diabetes, obesity, Parkinson’s, pain, and rehabilitation among others ^15–19^. This immersive technology is also being explored as a way to enhance mindfulness practice through VR-based apps incorporating aspects of mindfulness ^7,20^. A narrative review of seven VR apps for mindfulness which were trialled found VR-based mindfulness training seemed to be more effective than conventional mindfulness, helping to reduce anxiety, depression and improve sleep and mood and they can cause cyber sickness if a person remains immersed in the interactive audio-visual experience for too long ^21^. However, this review focused solely on scientifically developed VR apps for mindfulness and not commercially available ones.

While some VR-based mindfulness apps have been developed and evaluated by researchers, there are potentially other VR mindfulness apps available commercially via the online app stores that can be downloaded and used by patients and the public. Given the growing popularity of mindfulness practice and VR technologies a rigorous review of commercially available VR-based mindfulness apps that examines and reports their quality could add value for those considering using them. Furthermore, clear recommendations from commercial health app reviews can help improve the quality of future health apps ^22^ which could lead to better patient and health services outcomes long term. Hence, this review aimed to identify and evaluate commercially available VR-based apps for mindfulness.

## Methods

We undertook a commercial health app review following clear methodological guidance ^22^, and employed the Preferred Reporting Items for Systematic Reviews and Meta-Analyses (PRISMA) guidelines when conducting and reporting the review ^23^.

### Search strategy

We searched the major app stores (i.e., iOS (Apple app), Android (Google Play) and Meta) using relevant terms such as ‘mindfulness’, ‘mindfulness-meditation’, ‘VR mindfulness’, ‘VR mindfulness-meditation’, ‘VR mindfulness apps’, ‘VR mindfulness-based apps’, ‘VR mindfulness-based stress reduction apps’, ‘VR mindfulness-based cognitive therapy apps’, ‘VR mindfulness-meditation apps’ and ‘VR mental health apps’ between September and December 2023. The National Health Service (NHS) apps library (now closed) and the Organisation for the Review of Care and Health Apps (ORCHA) (https://orchahealth.com/our-products/health-app-library/) platforms were also searched with key terms.

The ‘Target user’ (T), ‘Evaluation’ (E), ‘Connectedness’ (C), and ‘Health domain’ (H) or ‘TECH’ framework was used to develop the eligibility criteria for the review ^22^. The target users were adults aged 18 years and above, excluding those aimed solely at children or adolescents. VR apps offered to both adults and young people were included as mindfulness can be practiced at any age. The evaluation focused on all types of VR apps (i.e., fully and partially immersive) that incorporated education, training, strategies, or techniques related to mindfulness. VR apps that only contained timers, reminders, or guided meditation were excluded as were online or web-based platforms offering mindfulness practice and standard mobile mindfulness apps as scientific reviews are published in these areas ^12,24,25^. VR apps that were under development or only available in a prototype form were also excluded. Connectedness centred on VR apps that did and did not connect to other devices or apps and we included both in the review. Finally, the Health domain was open ended to include all physical and mental health conditions as well as apps related to maintaining general health and wellbeing through practising mindfulness. VR apps were restricted to the English language.

### Screening and data extraction

We found two hundred and sixty-two VR apps related to mindfulness which we screened against the eligibility criteria and those not relevant were discarded. The remaining VR apps were downloaded to a Meta Quest 2 VR headset for further review and those not meeting the inclusion criteria were excluded. Any disagreements during screening were discussed among the research team to reach consensus (Figure 1, Appendix A). Next, relevant data were extracted from the VR app, the App store, and the website associated with the VR app to Microsoft Excel (e.g., app’s name, developer, version number, market/s in which it was available, cost to download and number of user ratings) (Tables 1 and 2).

**Figure 1.**
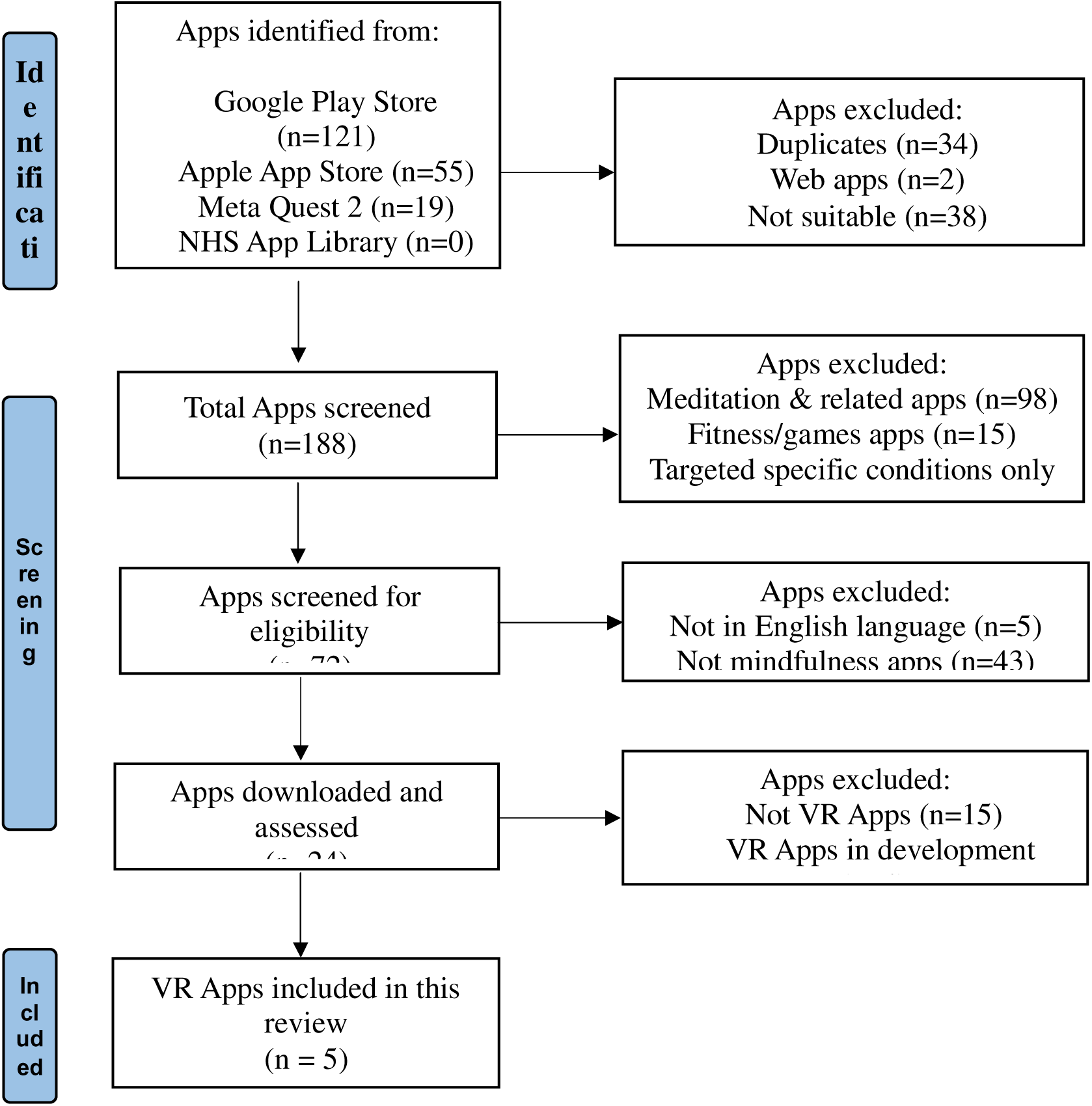
Prisma flow diagram to show selection of VR mindfulness apps

**Table 1.** Characteristics of VR apps related to mindfulness.

| App | Ages | Operating system | Supported VR platforms | Content | Viewer ratings | Number of viewer ratings | Developer | Download size | Release date |
| --- | --- | --- | --- | --- | --- | --- | --- | --- | --- |
| **Headspace XR** | All | Android & iOS | Meta Quest 2, 3 and Pro | Headspace is a virtual playground - provides an option to share the user's experience in social networks such as Facebook and Twitter. Headspace has an app community. Provides in-app purchase that includes additional guided meditation tracks. Internet connection not needed. https://www.headspace.com/xr | 4.4 | 44 | Nexus Studios | 1.94GB | 2024 |
| **Hoame** | All | Android/ Meta | Meta Quest 2, 3 and Pro | Has 350 guided meditations, nature landscapes, exercises, breath-work, and guided and self-guided meditation. Sound baths, visualizations, body-scans, or sleep classes. A 'salt cave' and 'infrared sauna' to promote restoration, relaxation, and mindfulness. Tranquil sandboxes. Daily study classes, events, workshops, logging of community progress, mood quizzes, customisation of meditation/mindfulness areas and 'on-demand' experiences. https://www.hoame.app/ | 3.3 | 249 | Hoame | 1.45GB | 2022 |
| **Innerworld** | 17+ | iOS 14.0 or later | Meta Quest, Meta Quest 2, 3 and Pro | Virtual community to explore behaviours, emotions, and thoughts through group support, events, and courses. Virtual guides run mental health meetings. Simple timers. https://www.inner.world/home/ | 4.5 | 482 | Innerworld | 255.43MB | 2022 |
| **Maloka** | All | Android 5.1 and up, and iOS | Meta Quest, Meta Quest 2, 3 and Pro | Users build their own tropical island which grows as they practice mindfulness. Accompanied by a 'spirit' guide or 'voice of the universe' who monitors and provides advice. Mindfulness library, Breathwork, Body scans, Sound baths, Guided meditation, Basic yoga. https://www.playmaloka.com/ | 4.2 | 471 | PlayMaloka | 922.62MB | 2021 |
| **TRIPP** | All | Android 10 and up, and iOS | Meta Quest, Meta Quest 2, 3 and Pro | Mood on demand (has a lifetime version), >100 guided immersive mindfulness/meditation exercises (short, medium, long sessions), sleep, mixed reality gifts, personalised experiences, and multi-day challenges. Offers visually guided breathing techniques, and mood logging. https://www.tripp.com/ | 4.2 | 3,000 | TRIPP Inc. | 2.33GB | 2019 |

**Table 2.** Additional characteristics of VR apps related to mindfulness.

| App | In-app purchases | Internet connection | Trial period/<br>free demo | Subscription availability | Privacy policy | App community | Social media | Guides/<br>Instructors | Mobile app version | Pricing |
| --- | --- | --- | --- | --- | --- | --- | --- | --- | --- | --- |
| **Headspace XR** | ? | ? | ? | ? | ? | ? | ? | ? | ? | Price: £22.99;<br>Internet connection not needed |
| **Hoame** | ? | ? | ?(7 days) | ? | ? | ? | ? | ? |  | 1 month pass: £7.99/month<br>(includes 1-week free trial);<br>6 month pass: £39.23/6 months;<br>1 year pass: £62.59/year<br>Internet connection needed |
| **Innerworld** | ? | ? | ? | ? | ? | ? | ? | ? | ? | Membership: £14.99/month<br>Membership: £99.99/year<br>Internet connection needed |
| **Maloka** | ? | ? | ? | ? | ? | ? | ? | ? | ? | Free on iOS, Google Play and<br>Meta Quest store;<br>Internet connection needed |
| **TRIPP** | ? | ? | ? | ? | ? | ? | ? | ? | ? | Membership: £7.99/month;<br>Membership: £29.99/year;<br>Internet connection needed |

### Analysis

Descriptive statistics were generated on key data to summarise aspects of the VR apps and we also used the Mobile App Rating Scale (MARS) to determine the quality of each VR app ^26^. MARS consists of 19 items across four dimensions (i.e., engagement, functionality, aesthetics and information quality), with each item rated on a 5-point Likert scale: (1) inadequate (2) poor (3) acceptable (4) good and (5) excellent. MARS has good reliability of the subscales (α = 0.80 to 0.89) and the overall scale (α = 0.90), as well as good objectivity (subscales: ICC = 0.50 to 0.80; overall = 0.90). In previous commercial health app reviews ^27,28^, the optional subscale for subjective quality was omitted to ensure that quality assessments were only objective. Six reviewers independently rated each VR mindfulness app using the MARS, with the mean scores per dimension and the mean overall quality score across dimensions calculated. Inter-rater reliability for the MARS scores was also assessed (Table 3, Appendix B), using SPSS Statistics (Version 29). Intra-class correlation coefficients (ICCs) were created on all MARS items to allow for the appropriate calculation of weighted values of rater agreement ^29^. Each reviewer also documented analytical memos for each VR app covering the four dimensions of MARS which were summarised descriptively. Finally, we searched for empirical evidence that evaluated the included VR apps to determine if their effectiveness, acceptability, usability or satisfaction had been scientifically evaluated.

**Table 3.** MARS mean quality scores of included VR mindfulness-based apps (all six raters)

| App | Engagement | Functionality | Aesthetics | Information | Overall | Subjective Quality |
| --- | --- | --- | --- | --- | --- | --- |
| Headspace XR | 3.34 | 3.59 | 3.45 | 3.07 | 3.37 | 2.29 |
| Hoame | 3.40 | 4.17 | 3.89 | 3.62 | 3.77 | 2.94 |
| Innerworld | 3.73 | 3.38 | 2.78 | 3.41 | 3.31 | 2.95 |
| Maloka | 3.87 | 3.55 | 3.55 | 3.43 | 3.61 | 3.00 |
| TRIPP | 4.07 | 4.22 | 4.28 | 3.67 | 4.06 | 3.72 |

## Results

### VR App Characteristics

Five VR apps related to mindfulness were included in the review, which were: 1) Headspace XR 2) Hoame 3) Innerworld 4) Maloka and 5) TRIPP, all released between 2019 and 2024 (Figure 1). The VR apps provided access to meditative and mindfulness sessions, guided by virtual instructors in some cases, and situated in a range of virtual landscapes accompanied by sound or music. The five VR apps were for all age groups except Innerworld, which started from 17 years of age (with parental guidance). All five VR apps were available on the Meta Quest 2, 3 and Pro VR platforms and ran on Android and iOS operating systems. The average user rating score was 4.12 out of 5 (range: 3.3 to 4.5) (Table 1). All five VR apps provided in-app purchases such as additional guided meditation tracks and reminders to practice mindfulness and other activities. Four VR apps required payment/subscriptions and one, Maloka, was free. The VR apps also allowed users to share their experiences via social media platforms such as Facebook/Twitter (Table 2).

### MARS Ratings

TRIPP received the highest average MARS score (4.06), followed by Hoame (3.77), Maloka (3.61), Headspace XR (3.37) and Innerworld (3.31) (Table 3). Some of the apps had high ratings on the MARS subscales of visual aesthetics, engagement, functionality and information quality (Table 3, Appendix B). The five included apps were evaluated by six raters and the intra-class correlation (ICC) indicated that agreement of raters was moderate (ICC = 0.72, 95% CI: 0.51 to 0.86) ^30^. The average overall quality rating for the 5 mindfulness-based apps reviewed was M=3.62 (SD=0.31, range 3.31–4.06), demonstrating moderate quality.

### Engagement

Engagement in the MARS checklist refers to whether an app is entertaining, interesting, customisable, interactive and the content (e.g., visual information, language and design) well targeted to users ^26^. All five VR apps offered different levels of engagement to increase user interest, with most rated as relatively entertaining. The average MARS ratings for engagement was M=3.68 (SD=0.39, range=3.34–4.07) (Table 3). For instance, TRIPP and Hoame offered tools such as a mood survey and progress tracker, while Maloka offered a virtual spirit guide, the ability to build a personalised virtual island, and provided users with rewards. Most of the VR apps provided settings that allowed users to customise features such as background music or sounds, avatars, content, notifications, prompts, reminders, and sharing options. Most VR apps also allowed a certain level of interaction with users being able to provide their mood and receive prompts and feedback. Hoame included add-on features like the “hoamie score” for a better experience and enabled users to capture and share images or videos. Hoame and TRIPP also allowed the presence of other VR app users in some of the meditative sessions although these shared virtual spaces could sometimes be empty. TRIPP and Innerworld offered live VR group activities and events, although some were not related to mindfulness practice while Maloka had a library of experiences users could share with their online community. Finally, the content in all five VR apps seemed appropriate for the target audience although Maloka, Innerworld and HeadSpace XR used animated or cartoonish graphics which may not appeal to adult audiences.

### Functionality

Functionality in the MARS checklist refers to the features (functions) and components (buttons, menus, icons) of an app, if it is easy to learn and navigate, and has a logical flow with gestural design. Most of the features of the five VR apps performed reasonably well with virtual environments and menus that were easy to use and logical to navigate with clear labels, icons and instructions. The average MARS ratings for functionality was M=3.78 (SD=0.56, range=3.38–4.22) (Table 3). However, it was not clear how to exit some virtual experiences and return to the main menu and the VR apps were sometimes slow or froze occasionally. Moving between screens or virtual settings tended to be logical and uninterrupted with smooth transitions, although audio options could be added in some cases to enable easier navigation. The gestural design features used included interactions such as taps, swipes or scrolls using the hand controllers which were relatively consistent and intuitive across most components or screens in the VR apps.

### Aesthetics

In the MARS checklist, aesthetics encompasses the graphic design, overall visual appeal of an app, its colour scheme and stylistic consistency. The layout of the VR apps in terms of the arrangement and size of buttons, icons, menus or content was satisfactory with simple and clear displays that looked professional. The average MARS ratings for aesthetics was M=3.59 (SD=0.24, range=2.78–4.28) (Table 3). The graphics were mostly high-quality, with Hoame in particular having high quality stereoscopic videos and photorealistic images. However, Innerworld’s and Maloka’s graphics were lower in quality with more animated or cartoonish virtual environments that appeared lower in resolution than the other VR apps. Overall, most of the VR apps were visually appealing with consistent colour schemes and patterns, shapes and virtual spaces scaled to represent realistic environments and experiences.

### Information

Finally, information in the MARS checklist refers to the quality of the information contained in the app (i.e., text, feedback, measures, references) and whether it was from a credible source. All VR apps matched how they were described on the App stores or the company website although the content was not always related to mindfulness practice with some apps more focused on meditation and wellbeing. The average MARS ratings for information was M=3.44 (SD=0.31, range=3.07–3.67) (Table 3). It was difficult to gauge the accuracy of information provided in the live virtual events and the expertise of those guiding virtual sessions, as well as claims made as to the effect of the VR app on a person’s physical and mental health. Some of the VR apps had specific and measurable goals such as users tracking their feelings, mood, or emotions and ranking themselves against others. The quality and quantity of information provided on the VR apps seemed appropriate and relevant to their stated aims, with different virtual worlds for users to experience and practice breathing and other meditative and relaxation techniques. Information in the VR apps was provided in a range of audio-visual and text formats in particular visual explanations of concepts through charts, graphs, images or videos.

## Discussion

Overall, the five commercially available VR apps related to mindfulness i.e., Headspace XR, Hoame, Innerworld, Maloka, and TRIPP had a moderate quality score (3.62), with the scores on functionality and engagement being high and TRIPP received the highest average MARS score. VR seems to offer a uniquely immersive experience that can deepen engagement with mindfulness practice and help overcome distractions, while being accessible and intuitive to users. VR environments in the apps reviewed also provided a calm environment to users that may make it easier to practice mindfulness, especially for a novice. Furthermore, simulated live sessions (e.g. within InnerWorld and TRIPP) offered additional virtual experiences that could be more authentic and personalised. While traditional meditation apps provide simplicity and convenience, making them suitable for a broad audience, they may lack the immersive benefits of VR ^31^.

As the MARS quality scores varied across the five VR apps, further research exploring how to design a VR app for mindfulness practice could add value. Only a few of the apps named people who they considered to be experts in the development of the VR environments or the delivery of the immersive and interactive content. Hence, it is not clear if end users were involved in co-designing these VR technologies. Using a range of co-design methods, tools and theories is becoming popular when creating digital mental health interventions as users can share their perspectives on the design features, functionality, and content they prefer, leading to more personalised and useable technologies ^32,33^. Involving experienced mindfulness practitioners and researchers in the design and development of virtual experiences and guidelines on when and how often to use a VR app for mindfulness practice in parallel with face-to-face individual or groups session could be beneficial for creating high-quality VR apps for mindfulness ^34^.

The five VR apps reviewed tended to educate, inform or remind users mainly for meditative practices, often using techniques such as exercise and relaxation. However, all five VR apps came from commercial sources which could limit their credibility and only two had some scientific evidence supporting them. An observational study examined the use of Innerworld in providing Cognitive Behavioural Immersion (CBI) to people (n=127) experiencing depression and anxiety, reporting those who used the VR app showed lower anxiety symptoms ^35^. A small pilot study also investigated the feasibility of using Innerworld for CBI with individuals (n=48) recovering from substance abuse disorder and found it increased their positive affect with participants liking the community approach, immersive experience, and anonymity provided by the VR app ^35^. A scoping review of VR apps in healthcare included Innerworld but only described how it works in terms of forming online health communities where peer-led events allow users to support each other with common health concerns, although the review did raise concerns over the potential impact of unqualified mental health support ^36^. TRIPP was tested in a small randomised controlled trial to assess its effect on quality of life, wellbeing and mood in cancer patients undergoing surgery (n=54). This study found no difference in quality of life between the intervention and control groups, although feelings and adherence rates improved in those using the VR app ^37^. TRIPP was also explored in a residential substance abuse treatment setting using focus groups with patients (n=25) who used the VR app and staff who provided support (n=11), revealing it seemed to help people engage with mindfulness practice to reduce negative emotions and cultivate a sense of wellbeing ^38^. While one study included Hoame when co-designing VR interventions with young people, it did not evaluate the VR app in any way ^39^ and no research studies exist for Headspace XR and Maloka. Overall, the scientific evidence base for all five VR apps is limited.

Although a systematic review of VR apps (n=7) for mindfulness training indicated they may improve mental health conditions in adult populations (e.g., levels of mindfulness, meditation, sleep quality, emotion regulation), the quality of the VR apps were not evaluated, the VR training was limited to single sessions in some cases, and standardised measures to examine mindfulness and meditation-related outcomes were not always used ^21^. Hence, robust research investigating their feasibility and effectiveness on physical and mental health outcomes of different adult and child populations is needed ^14^. Furthermore, while the MARS tool is a useful checklist to assess the quality of a mobile health app, it does not cover all features of digital tools such as their privacy and security. Martinez-Perez et al. ^40^ highlight that many health apps do not have enough security and privacy mechanisms to protect user data, something all VR users should be aware of particularly when providing personal information on VR apps as it may be shared with others.

Along with a limited scientific evidence base, VR apps have additional challenges. First, they need to be used safely as a small number of participants may experience harm during mindfulness practice e.g., being re-traumatised. An experienced practitioner could reduce the likelihood of this occurring by assessing risk beforehand and adapting the delivery and content of the mindfulness session ^41^. In a VR app this risk could be mitigated by using a disclaimer clearly stating it should not be used a therapeutic tool and recommending users access a qualified healthcare practitioner if they have mental health concerns. Hence, VR apps could be used as supportive tools alongside face-to-face mindfulness practice so those who require trauma-informed and neurodiversity-informed adaptations can be catered for. Traditional MBPs are designed to unfold gently over time so that mindfulness teachers can reassure novices of the practices to follow, help dispel myths about mindfulness and support the process of personal inquiry and reflection helping to shape and improve mindfulness practice. These nuanced in person discussions and personal connections may not easily be replicated in a VR environment which may be best placed as an adjunct to traditional MBPs. However, for those who are uncomfortable with in-person group sessions, VR may present a useful way to widen access to mindfulness practice.

Second, VR headsets and hand controllers are not always easy and comfortable to use and could cause discomfort if worn or used incorrectly ^42^. The software may also be hard to use and the three dimensional virtual spaces difficult to navigate, particularly if someone’s digital literacy skills are low ^19^. If used for too long a period, a person may also experience VR-induced side effects such as eyestrain, nausea, dizziness, headaches, and mental overload ^43,44^. Therefore, clear guidelines outlining when, how often, and for how long a person should use a VR app to practice mindfulness could be provided along with hands-on training for those that need it. Third, many VR apps cost money and users need to subscribe to a weekly, monthly or annual fee, along with having access to the Internet to enable content and updates to be downloaded to a VR headset. Fourth, the VR equipment also needs to be charged on a regular basis so the battery life can sustain periods of use, with some devices needing to be replaced or upgraded after a few years which brings additional costs ^45^. These barriers may prevent some groups of people such as those from lower socioeconomic groups or with declining visual acuity or cognitive function from using a VR app ^46^. Thus, people could be encouraged to use freely available apps developed by mindfulness researchers or practitioners instead of commercially developed ones that require one off or regular payments, or use low fidelity (i.e., cardboard) VR headsets and standard smartphone/mobile apps for mindfulness practice which are less battery intensive.

## Strengths and limitations

This is the first commercial health app review to formally identify VR apps related to mindfulness and assess the quality of engagement, functionality, aesthetics and information. We followed clear methodological guidelines to conduct the review ^22^ and a validated, multi-dimensional measurement instrument, i.e., MARS, was used by six raters (including three experienced mindfulness practitioners and researchers) to assess the quality of the VR apps^26^. However, the review has a number of limitations. Although an extensive search of the VR App stores was conducted, only apps available in the UK and in English were included. Therefore, VR apps related to mindfulness that were released privately, available in other countries or in other languages, may have been missed, which limits the generalisability of the results. In addition, the MARS tool was developed for standard mobile health apps used on smartphones and not for VR based apps that require VR headsets and hand controllers. As these technologies have additional design features and functionality, the MARS tool may not have captured all aspects of the VR apps which may limit the utility of the review findings.

## Conclusion

The use of VR for mindfulness practice is in an early stage of development and this review provides important insights into VR apps related to mindfulness such as their availability and quality (i.e., engagement, functionality, aesthetics and information). Only five VR apps i.e., Headspace XR, Hoame, Innerworld, Maloka, and TRIPP were identified related to mindfulness practice with an overall moderate MARS quality score (3.62/5.00). TRIPP received the highest average MARS score (3.89), followed by Hoame (3.62), Maloka (3.53), Innerworld (3.38), and Headspace XR (3.34). These VR apps may provide a convenient and immersive way to access and engage in regular mindfulness practice, particularly for novices, and may address some of the limitations with face-to-face and mobile app mindfulness programmes. However, rigorous scientific research is needed to assess effectiveness of these VR apps in improving physical and mental health outcomes through immersive mindfulness practice.

## Ethical approval

Not applicable.

## Abbreviations

ICC – Intraclass correlation coefficient; MARS - Mobile App Rating Scale; MBPs - Mindfulness based programmes; NHS - National Health Service; ORCHA - Organisation for the Review of Care and Health Apps; PRISMA - Preferred Reporting Items for Systematic Reviews and Meta-Analyses; SPSS - Statistical Package for the Social Sciences; TECH - Target audience, Evaluation, Connectedness, Health domain; UK - United Kingdom; VR - Virtual Reality

## Acknowledgements

None.

## Data availability

All data produced in the present study are available upon reasonable request to the authors.

## CRediT authorship contribution statement

**Shraboni Ghosal:** Methodology, Data curation, Investigation, Formal analysis, Writing – original draft, **Mengying Zhang:** Methodology, Formal analysis, Writing – review & editing. **Angeliki Bogosian:** Methodology, Formal analysis, Writing – review & editing. **Elizabeth Marsh:** Methodology, Formal analysis, Writing – review & editing. **Trudi Edginton:** Methodology, Formal analysis, Writing – review & editing. **Emma Stanmore:** Funding acquisition, Methodology, Investigation, Formal analysis, Supervision, Writing – review & editing. **Siobhan O’Connor:** Conceptualisation, Funding acquisition, Methodology, Investigation, Formal analysis, Supervision, Writing – review & editing.

## Appendix A. List of applications excluded from the review

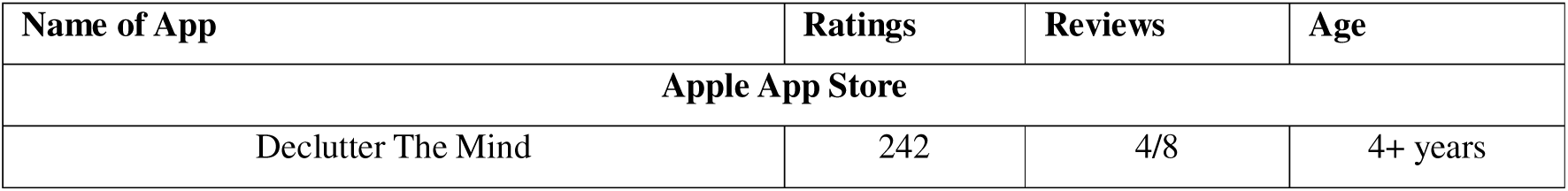
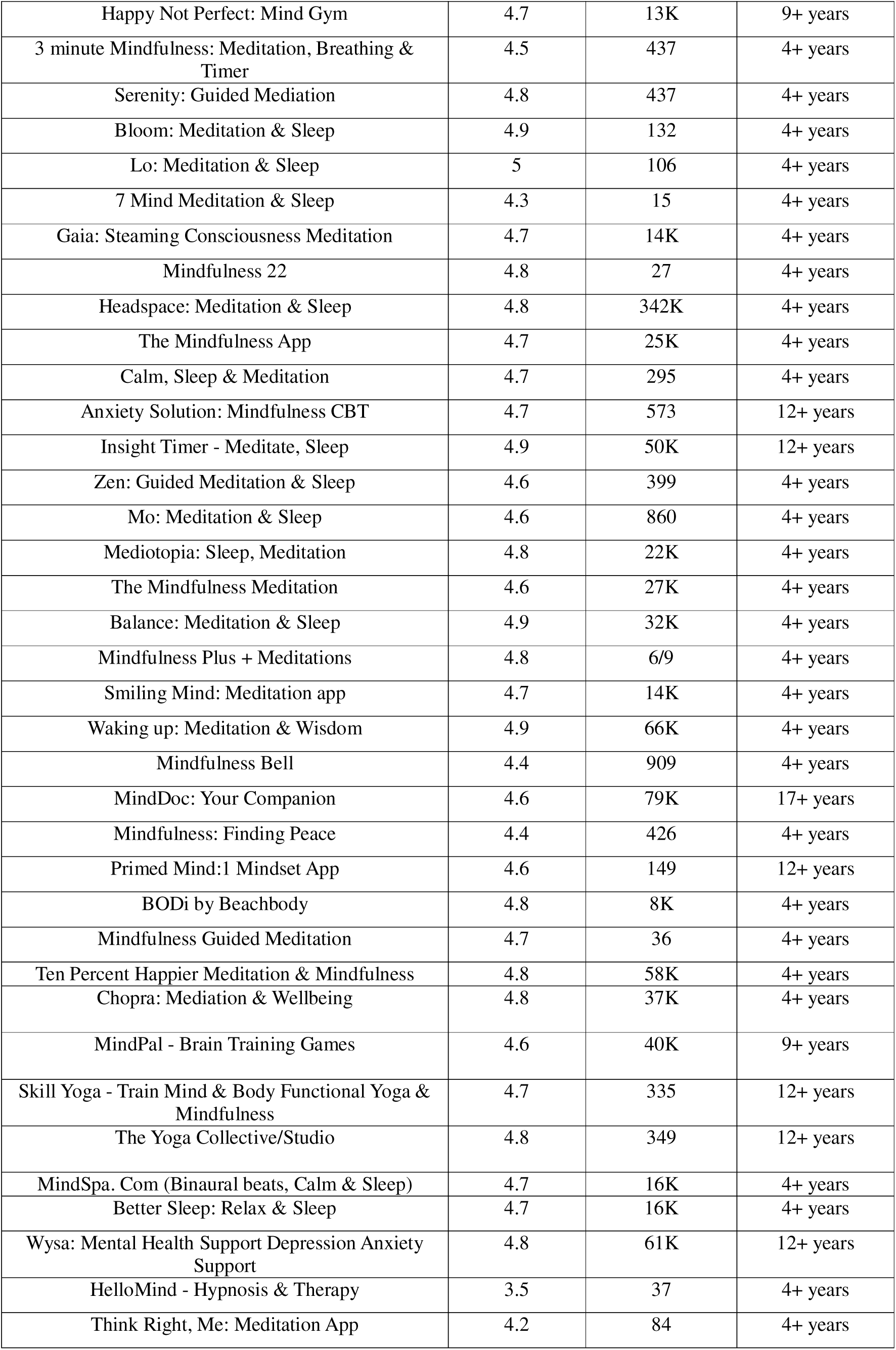
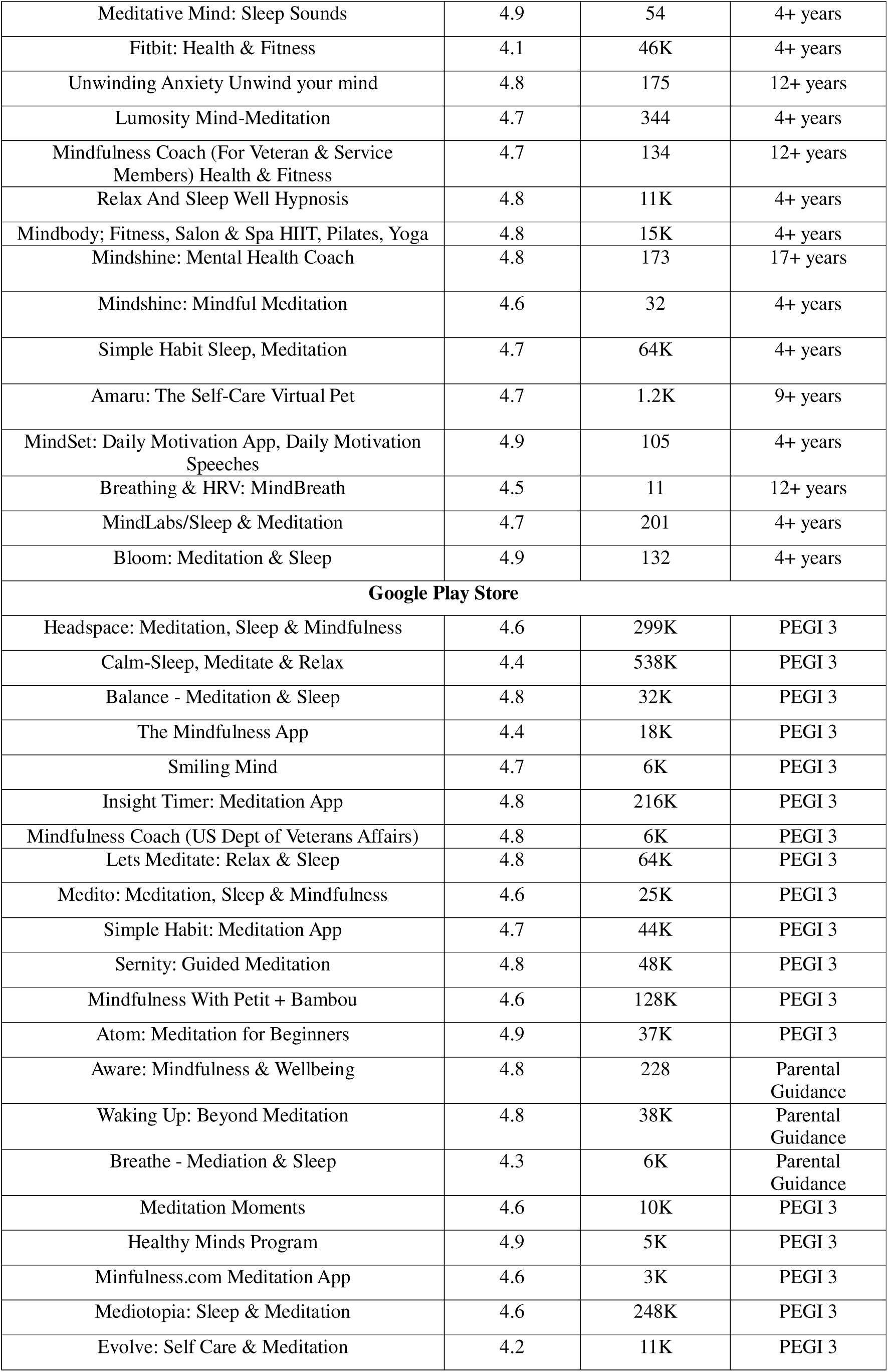
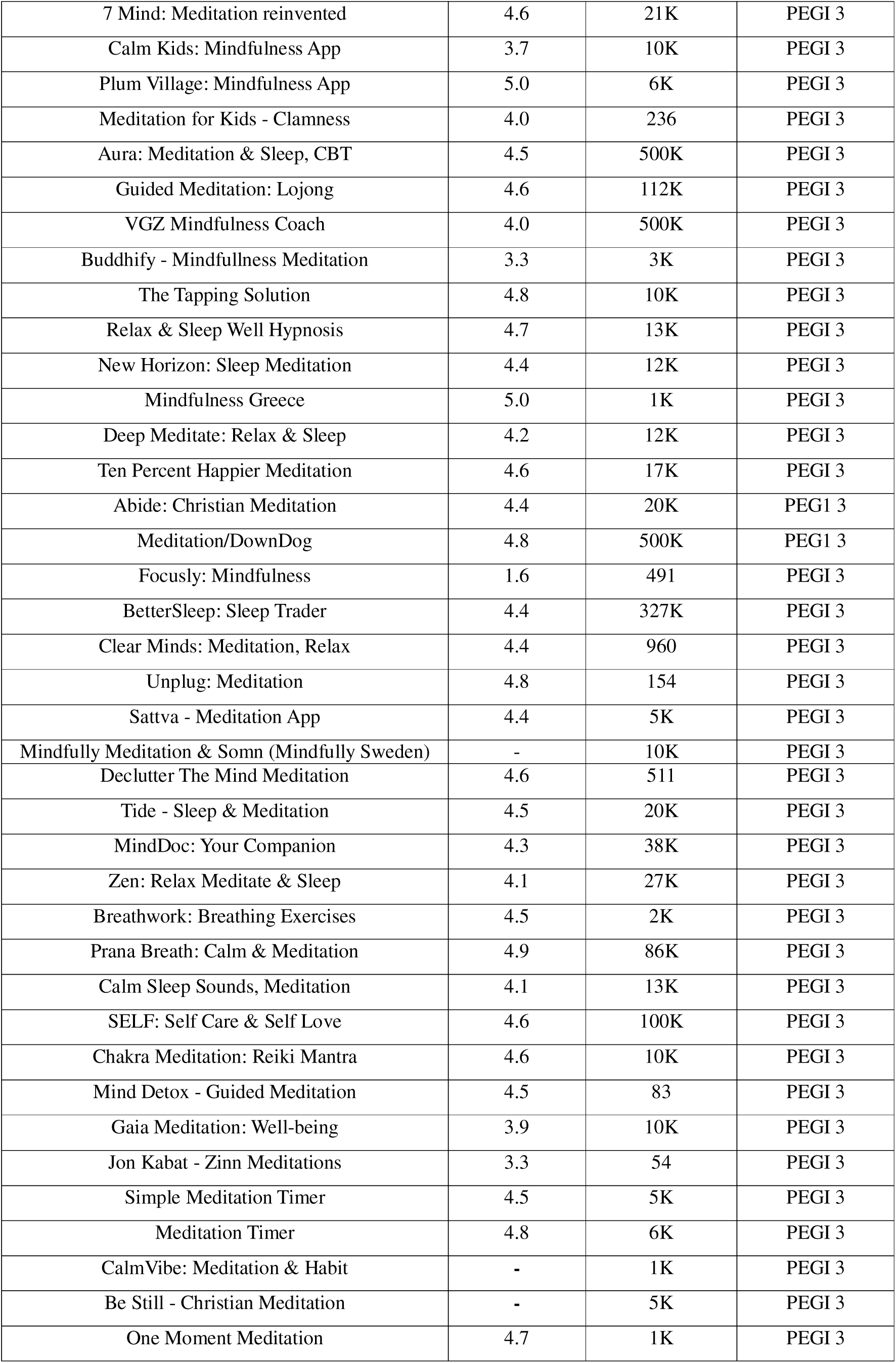
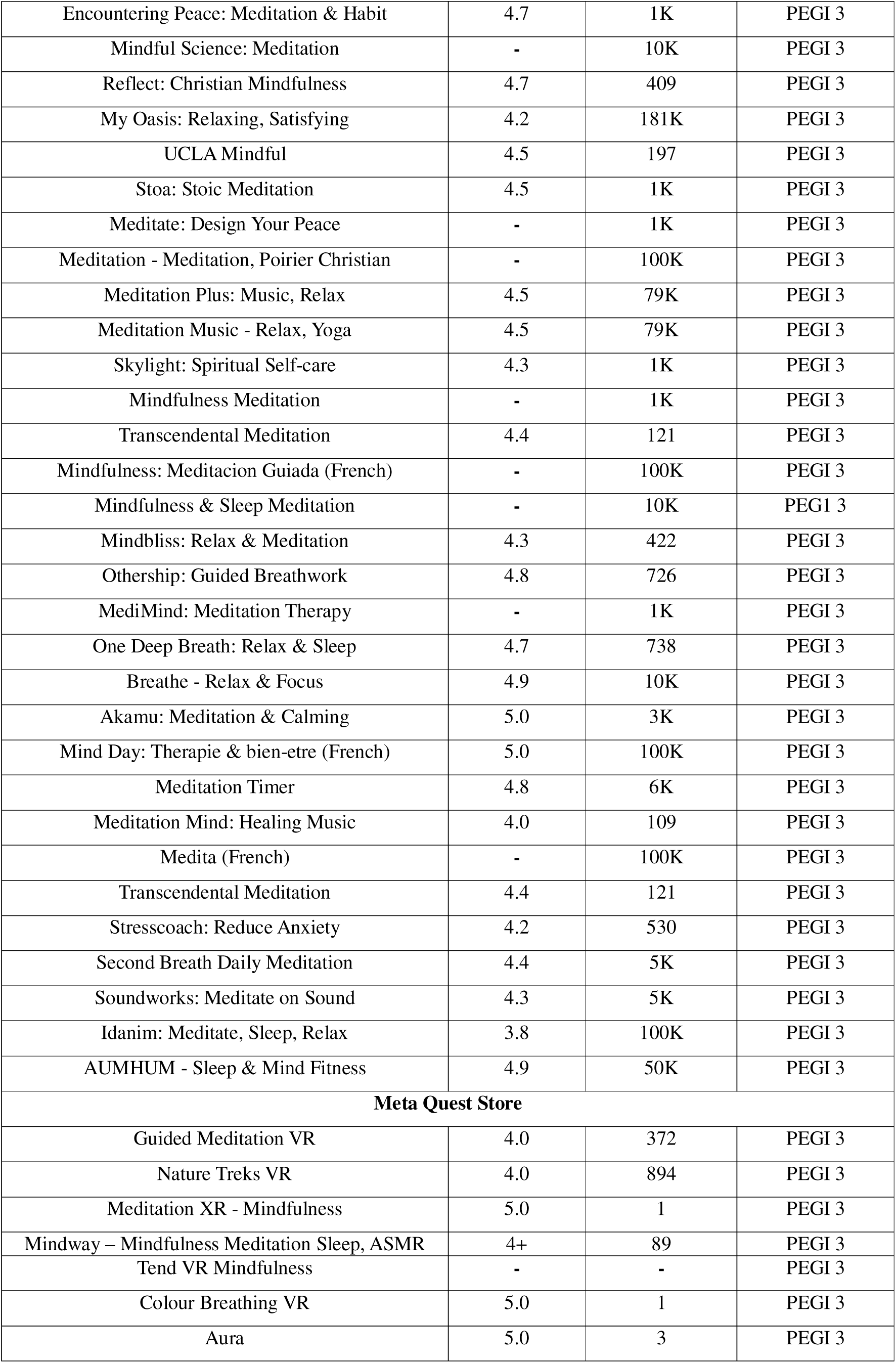
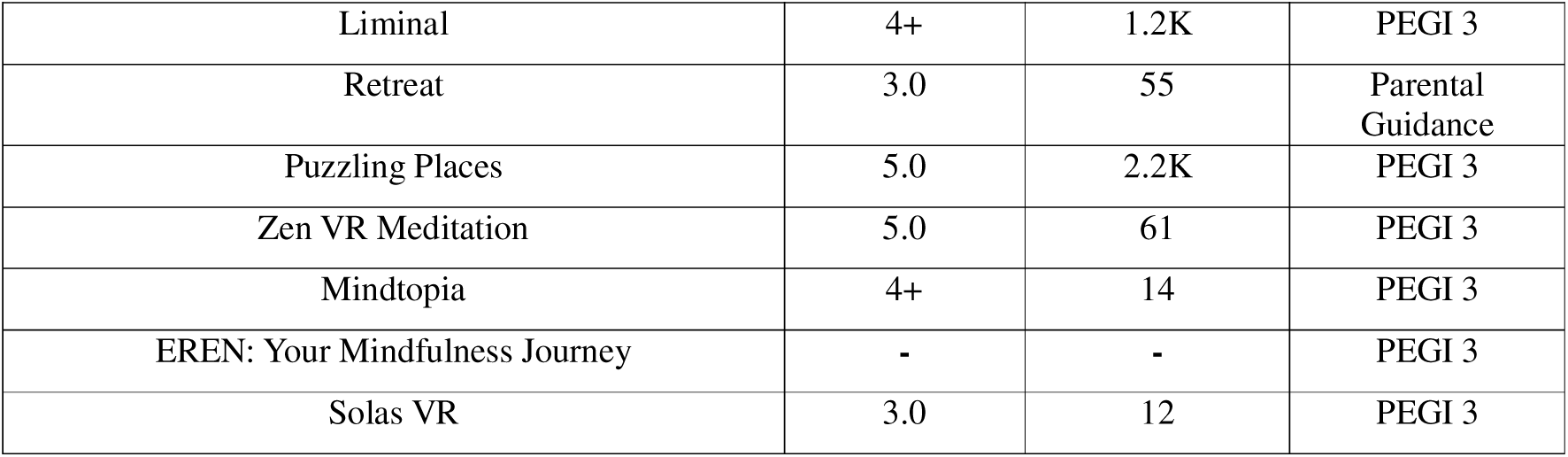

| Name of App | Ratings | Reviews | Age |
| --- | --- | --- | --- |
| **Apple App Store** |  |  |  |
| Declutter The Mind | 242 | 4/8 | 4+ years |
| Happy Not Perfect: Mind Gym | 4.7 | 13K | 9+ years |
| 3 minute Mindfulness: Meditation, Breathing & Timer | 4.5 | 437 | 4+ years |
| Serenity: Guided Meditation | 4.8 | 437 | 4+ years |
| Bloom: Meditation & Sleep | 4.9 | 132 | 4+ years |
| Lo: Meditation & Sleep | 5 | 106 | 4+ years |
| 7 Mind Meditation & Sleep | 4.3 | 15 | 4+ years |
| Gaia: Steaming Consciousness Meditation | 4.7 | 14K | 4+ years |
| Mindfulness 22 | 4.8 | 27 | 4+ years |
| Headspace: Meditation & Sleep | 4.8 | 342K | 4+ years |
| The Mindfulness App | 4.7 | 25K | 4+ years |
| Calm, Sleep & Meditation | 4.7 | 295 | 4+ years |
| Anxiety Solution: Mindfulness CBT | 4.7 | 573 | 12+ years |
| Insight Timer - Meditate, Sleep | 4.9 | 50K | 12+ years |
| Zen: Guided Meditation & Sleep | 4.6 | 399 | 4+ years |
| Mo: Meditation & Sleep | 4.6 | 860 | 4+ years |
| Mediotopia: Sleep, Meditation | 4.8 | 22K | 4+ years |
| The Mindfulness Meditation | 4.6 | 27K | 4+ years |
| Balance: Meditation & Sleep | 4.9 | 32K | 4+ years |
| Mindfulness Plus + Meditations | 4.8 | 6/9 | 4+ years |
| Smiling Mind: Meditation app | 4.7 | 14K | 4+ years |
| Waking up: Meditation & Wisdom | 4.9 | 66K | 4+ years |
| Mindfulness Bell | 4.4 | 909 | 4+ years |
| MindDoc: Your Companion | 4.6 | 79K | 17+ years |
| Mindfulness: Finding Peace | 4.4 | 426 | 4+ years |
| Primed Mind:1 Mindset App | 4.6 | 149 | 12+ years |
| BODi by Beachbody | 4.8 | 8K | 4+ years |
| Mindfulness Guided Meditation | 4.7 | 36 | 4+ years |
| Ten Percent Happier Meditation & Mindfulness | 4.8 | 58K | 4+ years |
| Chopra: Meditation & Wellbeing | 4.8 | 37K | 4+ years |
| MindPal - Brain Training Games | 4.6 | 40K | 9+ years |
| Skill Yoga - Train Mind & Body Functional Yoga & Mindfulness | 4.7 | 335 | 12+ years |
| The Yoga Collective/Studio | 4.8 | 349 | 12+ years |
| MindSpa. Com (Binaural beats, Calm & Sleep) | 4.7 | 16K | 4+ years |
| Better Sleep: Relax & Sleep | 4.7 | 16K | 4+ years |
| Wysa: Mental Health Support Depression Anxiety Support | 4.8 | 61K | 12+ years |
| HelloMind - Hypnosis & Therapy | 3.5 | 37 | 4+ years |
| Think Right, Me: Meditation App | 4.2 | 84 | 4+ years |
| Meditative Mind: Sleep Sounds | 4.9 | 54 | 4+ years |
| Fitbit: Health & Fitness | 4.1 | 46K | 4+ years |
| Unwinding Anxiety Unwind your mind | 4.8 | 175 | 12+ years |
| Lumosity Mind-Meditation | 4.7 | 344 | 4+ years |
| Mindfulness Coach (For Veteran & Service Members) Health & Fitness | 4.7 | 134 | 12+ years |
| Relax And Sleep Well Hypnosis | 4.8 | 11K | 4+ years |
| Mindbody: Fitness, Salon & Spa HIIT, Pilates, Yoga | 4.8 | 15K | 4+ years |
| Mindshine: Mental Health Coach | 4.8 | 173 | 17+ years |
| Mindshine: Mindful Meditation | 4.6 | 32 | 4+ years |
| Simple Habit Sleep, Meditation | 4.7 | 64K | 4+ years |
| Amaru: The Self-Care Virtual Pet | 4.7 | 1.2K | 9+ years |
| MindSet: Daily Motivation App, Daily Motivation Speeches | 4.9 | 105 | 4+ years |
| Breathing & HRV: MindBreath | 4.5 | 11 | 12+ years |
| MindLabs/Sleep & Meditation | 4.7 | 201 | 4+ years |
| Bloom: Meditation & Sleep | 4.9 | 132 | 4+ years |
| **Google Play Store** |  |  |  |
| Headspace: Meditation, Sleep & Mindfulness | 4.6 | 299K | PEGI 3 |
| Calm-Sleep, Meditate & Relax | 4.4 | 538K | PEGI 3 |
| Balance - Meditation & Sleep | 4.8 | 32K | PEGI 3 |
| The Mindfulness App | 4.4 | 18K | PEGI 3 |
| Smiling Mind | 4.7 | 6K | PEGI 3 |
| Insight Timer: Meditation App | 4.8 | 216K | PEGI 3 |
| Mindfulness Coach (US Dept of Veterans Affairs) | 4.8 | 6K | PEGI 3 |
| Lets Meditate: Relax & Sleep | 4.8 | 64K | PEGI 3 |
| Medito: Meditation, Sleep & Mindfulness | 4.6 | 25K | PEGI 3 |
| Simple Habit: Meditation App | 4.7 | 44K | PEGI 3 |
| Serinity: Guided Meditation | 4.8 | 48K | PEGI 3 |
| Mindfulness With Petit + Bambou | 4.6 | 128K | PEGI 3 |
| Atom: Meditation for Beginners | 4.9 | 37K | PEGI 3 |
| Aware: Mindfulness & Wellbeing | 4.8 | 228 | Parental Guidance |
| Waking Up: Beyond Meditation | 4.8 | 38K | Parental Guidance |
| Breathe - Meditation & Sleep | 4.3 | 6K | Parental Guidance |
| Meditation Moments | 4.6 | 10K | PEGI 3 |
| Healthy Minds Program | 4.9 | 5K | PEGI 3 |
| Minfulness.com Meditation App | 4.6 | 3K | PEGI 3 |
| Mediotopia: Sleep & Meditation | 4.6 | 248K | PEGI 3 |
| Evolve: Self Care & Meditation | 4.2 | 11K | PEGI 3 |
| 7 Mind: Meditation reinvented | 4.6 | 21K | PEGI 3 |
| Calm Kids: Mindfulness App | 3.7 | 10K | PEGI 3 |
| Plum Village: Mindfulness App | 5.0 | 6K | PEGI 3 |
| Meditation for Kids - Clamness | 4.0 | 236 | PEGI 3 |
| Aura: Meditation & Sleep, CBT | 4.5 | 500K | PEGI 3 |
| Guided Meditation: Lojong | 4.6 | 112K | PEGI 3 |
| VGZ Mindfulness Coach | 4.0 | 500K | PEGI 3 |
| Buddhify - Mindfulness Meditation | 3.3 | 3K | PEGI 3 |
| The Tapping Solution | 4.8 | 10K | PEGI 3 |
| Relax & Sleep Well Hypnosis | 4.7 | 13K | PEGI 3 |
| New Horizon: Sleep Meditation | 4.4 | 12K | PEGI 3 |
| Mindfulness Greece | 5.0 | 1K | PEGI 3 |
| Deep Meditate: Relax & Sleep | 4.2 | 12K | PEGI 3 |
| Ten Percent Happier Meditation | 4.6 | 17K | PEGI 3 |
| Abide: Christian Meditation | 4.4 | 20K | PEGI 3 |
| Meditation/DownDog | 4.8 | 500K | PEGI 3 |
| Focusly: Mindfulness | 1.6 | 491 | PEGI 3 |
| BetterSleep: Sleep Trader | 4.4 | 327K | PEGI 3 |
| Clear Minds: Meditation, Relax | 4.4 | 960 | PEGI 3 |
| Unplug: Meditation | 4.8 | 154 | PEGI 3 |
| Sattva - Meditation App | 4.4 | 5K | PEGI 3 |
| Mindfully Meditation & Somn (Mindfully Sweden) | - | 10K | PEGI 3 |
| Declutter The Mind Meditation | 4.6 | 511 | PEGI 3 |
| Tide - Sleep & Meditation | 4.5 | 20K | PEGI 3 |
| MindDoc: Your Companion | 4.3 | 38K | PEGI 3 |
| Zen: Relax Meditate & Sleep | 4.1 | 27K | PEGI 3 |
| Breathwork: Breathing Exercises | 4.5 | 2K | PEGI 3 |
| Prana Breath: Calm & Meditation | 4.9 | 86K | PEGI 3 |
| Calm Sleep Sounds, Meditation | 4.1 | 13K | PEGI 3 |
| SELF: Self Care & Self Love | 4.6 | 100K | PEGI 3 |
| Chakra Meditation: Reiki Mantra | 4.6 | 10K | PEGI 3 |
| Mind Detox - Guided Meditation | 4.5 | 83 | PEGI 3 |
| Gaia Meditation: Well-being | 3.9 | 10K | PEGI 3 |
| Jon Kabat - Zinn Meditations | 3.3 | 54 | PEGI 3 |
| Simple Meditation Timer | 4.5 | 5K | PEGI 3 |
| Meditation Timer | 4.8 | 6K | PEGI 3 |
| CalmVibe: Meditation & Habit | - | 1K | PEGI 3 |
| Be Still - Christian Meditation | - | 5K | PEGI 3 |
| One Moment Meditation | 4.7 | 1K | PEGI 3 |
| Encountering Peace: Meditation & Habit | 4.7 | 1K | PEGI 3 |
| Mindful Science: Meditation | - | 10K | PEGI 3 |
| Reflect: Christian Mindfulness | 4.7 | 409 | PEGI 3 |
| My Oasis: Relaxing, Satisfying | 4.2 | 181K | PEGI 3 |
| UCLA Mindful | 4.5 | 197 | PEGI 3 |
| Stoa: Stoic Meditation | 4.5 | 1K | PEGI 3 |
| Meditate: Design Your Peace | - | 1K | PEGI 3 |
| Meditation - Meditation, Poirier Christian | - | 100K | PEGI 3 |
| Meditation Plus: Music, Relax | 4.5 | 79K | PEGI 3 |
| Meditation Music - Relax, Yoga | 4.5 | 79K | PEGI 3 |
| Skylight: Spiritual Self-care | 4.3 | 1K | PEGI 3 |
| Mindfulness Meditation | - | 1K | PEGI 3 |
| Transcendental Meditation | 4.4 | 121 | PEGI 3 |
| Mindfulness: Meditacion Guiada (French) | - | 100K | PEGI 3 |
| Mindfulness & Sleep Meditation | - | 10K | PEGI 3 |
| Mindbliss: Relax & Meditation | 4.3 | 422 | PEGI 3 |
| Othership: Guided Breathwork | 4.8 | 726 | PEGI 3 |
| MediMind: Meditation Therapy | - | 1K | PEGI 3 |
| One Deep Breath: Relax & Sleep | 4.7 | 738 | PEGI 3 |
| Breathe - Relax & Focus | 4.9 | 10K | PEGI 3 |
| Akamu: Meditation & Calming | 5.0 | 3K | PEGI 3 |
| Mind Day: Therapie & bien-etre (French) | 5.0 | 100K | PEGI 3 |
| Meditation Timer | 4.8 | 6K | PEGI 3 |
| Meditation Mind: Healing Music | 4.0 | 109 | PEGI 3 |
| Medita (French) | - | 100K | PEGI 3 |
| Transcendental Meditation | 4.4 | 121 | PEGI 3 |
| Stresscoach: Reduce Anxiety | 4.2 | 530 | PEGI 3 |
| Second Breath Daily Meditation | 4.4 | 5K | PEGI 3 |
| Soundworks: Meditate on Sound | 4.3 | 5K | PEGI 3 |
| Idanim: Meditate, Sleep, Relax | 3.8 | 100K | PEGI 3 |
| AUMHUM - Sleep & Mind Fitness | 4.9 | 50K | PEGI 3 |
| **Meta Quest Store** |  |  |  |
| Guided Meditation VR | 4.0 | 372 | PEGI 3 |
| Nature Treks VR | 4.0 | 894 | PEGI 3 |
| Meditation XR - Mindfulness | 5.0 | 1 | PEGI 3 |
| Mindway – Mindfulness Meditation Sleep, ASMR | 4+ | 89 | PEGI 3 |
| Tend VR Mindfulness | - | - | PEGI 3 |
| Colour Breathing VR | 5.0 | 1 | PEGI 3 |
| Aura | 5.0 | 3 | PEGI 3 |
| Liminal | 4+ | 1.2K | PEGI 3 |
| Retreat | 3.0 | 55 | Parental Guidance |
| Puzzling Places | 5.0 | 2.2K | PEGI 3 |
| Zen VR Meditation | 5.0 | 61 | PEGI 3 |
| Mindtopia | 4+ | 14 | PEGI 3 |
| EREN: Your Mindfulness Journey | - | - | PEGI 3 |
| Solas VR | 3.0 | 12 | PEGI 3 |

## Appendix B: MARS quality scores of the included VR mindfulness-based apps by six raters

**B.1** MARS quality scores of the included VR mindfulness-based apps (1^st^ rater - SG)

Appendix B: MARS quality scores of the included VR mindfulness-based apps by six raters B.1 MARS quality scores of the included VR mindfulness-based apps (1<sup>st</sup> rater - SG)
| App | Engagement | Functionality | Aesthetics | Information | Overall | Subjective Quality |
| --- | --- | --- | --- | --- | --- | --- |
| Headspace XR | 4.00 | 4.00 | 4.00 | 4.00 | 4.00 | 4.25 |
| Hoame | 4.00 | 3.75 | 3.66 | 3.70 | 3.78 | 3.75 |
| Innerworld | 4.00 | 4.00 | 3.66 | 3.57 | 3.81 | 3.75 |
| Maloka | 4.00 | 4.00 | 3.66 | 3.42 | 3.77 | 3.75 |
| TRIPP | 4.00 | 4.00 | 4.00 | 3.71 | 3.93 | 4.25 |

**B.2** MARS quality scores of the included VR mindfulness-based apps (2^nd^ rater - SOC)

| App | Engagement | Functionality | Aesthetics | Information | Overall | Subjective Quality |
| --- | --- | --- | --- | --- | --- | --- |
| Headspace XR | 3.00 | 3.75 | 3.33 | 2.67 | 3.19 | 1.75 |
| Hoame | 3.40 | 4.00 | 3.66 | 3.17 | 3.56 | 2.25 |
| Innerworld | 2.40 | 2.50 | 2.66 | 2.33 | 2.47 | 1.25 |
| Maloka | 3.60 | 3.75 | 3.33 | 3.00 | 3.42 | 2.75 |
| TRIPP | 3.60 | 4.00 | 4.00 | 3.17 | 3.69 | 2.75 |

**B.3** MARS quality scores of the included VR mindfulness-based apps (3^rd^ rater - EM)

| App | Engagement | Functionality | Aesthetics | Information | Overall | Subjective Quality |
| --- | --- | --- | --- | --- | --- | --- |
| Headspace XR | 4.20 | 3.80 | 3.30 | 2.90 | 3.60 | 2.50 |
| Hoame | 3.60 | 4.00 | 3.30 | 2.90 | 3.50 | 2.30 |
| Innerworld | 4.20 | 3.00 | 2.30 | 2.90 | 3.10 | 1.25 |
| Maloka | 3.80 | 3.50 | 3.00 | 2.40 | 3.20 | 2.50 |
| TRIPP | 4.40 | 3.30 | 4.70 | 2.60 | 3.75 | 3.00 |

**B.4** MARS quality scores of the included VR mindfulness-based apps (4^th^ rater - AB)

| App | Engagement | Functionality | Aesthetics | Information | Overall | Subjective Quality |
| --- | --- | --- | --- | --- | --- | --- |
| Headspace XR | 3.25 | 3.25 | 3.70 | 2.60 | 3.20 | 1.75 |
| Hoame | 2.20 | 4.00 | 3.66 | 3.00 | 3.20 | 2.75 |
| Innerworld | 4.20 | 3.00 | 2.66 | 4.00 | 3.47 | 4.00 |
| Maloka | 3.00 | 3.25 | 3.33 | 3.20 | 3.31 | 2.25 |
| TRIPP | 4.00 | 4.50 | 4.00 | 4.00 | 4.13 | 4.00 |

**B.5** MARS quality scores of the included VR mindfulness-based apps (5^th^ rater - MZ)

| App | Engagement | Functionality | Aesthetics | Information | Overall | Subjective Quality |
| --- | --- | --- | --- | --- | --- | --- |
| Headspace XR | 1.80 | 3.25 | 2.67 | 3.17 | 2.72 | 1.50 |
| Hoame | 3.40 | 4.25 | 4.33 | 4.17 | 4.04 | 3.33 |
| Innerworld | 4.60 | 4.00 | 3.67 | 3.86 | 4.03 | 4.67 |
| Maloka | 4.20 | 3.75 | 4.00 | 3.83 | 3.95 | 4.00 |
| TRIPP | 3.40 | 4.50 | 4.00 | 3.83 | 3.93 | 3.33 |

**B.6** MARS quality scores of the included VR mindfulness-based apps (6^th^ rater - TE)

| **App** | **Engagement** | **Functionality** | **Aesthetics** | **Information** | **Overall** | **Subjective Quality** |
| --- | --- | --- | --- | --- | --- | --- |
| **Headspace XR** | 3.80 | 3.50 | 3.70 | 3.10 | 3.52 | 2.00 |
| **Hoame** | 3.80 | 5.00 | 4.70 | 4.70 | 4.55 | 3.25 |
| **Innerworld** | 3.00 | 3.75 | 1.70 | 3.80 | 3.00 | 2.75 |
| **Maloka** | 4.60 | 3.00 | 4.00 | 4.70 | 4.00 | 2.75 |
| **TRIPP** | 5.00 | 5.00 | 5.00 | 4.70 | 4.90 | 5.00 |

